# Comparison of gamma-aminobutyric acid, glutamate, and N-acetylaspartate concentrations in the insular cortex between patients with fibromyalgia, rheumatoid arthritis, and healthy controls - a magnetic resonance spectroscopy study

**DOI:** 10.1101/2022.09.15.22279963

**Authors:** Hans-Christoph Aster, Viola Hahn, Marc Schmalzing, György A. Homola, Thomas Kampf, Mirko Pham, Nurcan Üçeyler, Claudia Sommer

**Affiliations:** Department of Neurology, University Hospital Würzburg; Department of Neuroradiology, University Hospital Würzburg; Rheumatology/Clinical Immunology, Department of Internal Medicine II, University Hospital Würzburg

## Abstract

Fibromyalgia syndrome (FMS) is a chronic pain disorder with hypersensitivity to painful stimuli. A subgroup of patients shows damage to small peripheral nerve fibers. Previous studies support the hypothesis that increased activation of the pain-processing insular cortex is mediated by an imbalance of insular glutamate and γ-aminobutyric acid (GABA) concentrations. Here, we aimed to test this hypothesis in a large cohort of FMS patients comparing data of patients and healthy controls. In addition, we tested the hypothesis whether a reduction in small peripheral nerve fibers relates to glutamate concentrations in the insular cortex. We recruited 102 subjects (all female, 44 FMS patients, 40 healthy age-matched controls, and 19 patients with rheumatoid arthritis (RA) as disease controls. Study participants underwent single-voxel magnetic resonance spectroscopy of the right and left insular cortex. All patients completed questionnaires on symptom severity (pain intensity, impairment due to symptoms, depression). FMS patients were further stratified into subgroups with and without reduced intraepidermal nerve fiber density (IENFD) assessed on skin punch biopsies. We found no intergroup difference of the glutamate/GABA metabolite concentrations between FMS and RA patients and healthy controls. Glutamate/GABA levels did not correlate with symptom severity. Cerebral glutamate concentrations were independent of skin innervation. We found similar insular glutamate/GABA concentrations in FMS patients and disease and healthy controls. Therefore, our data cannot support the hypothesis that a glutamate/GABA mismatch leads to a sensitization of the insular cortex of fibromyalgia patients and thereby induces the symptoms.

## Introduction

Patients with fibromyalgia syndrome (FMS) suffer from widespread pain and comorbid symptoms like depression and anxiety [1]. FMS prevalence ranges from 2-4%, with more women affected [2]. The pathophysiology of FMS is not fully understood. Currently, a multifactorial origin including alterations in the peripheral [3, 4], autonomic [5], and central nervous systems (CNS) [6] is assumed. Genetic [7], hormonal [8], and psychosocial factors such as childhood trauma may facilitate the development of FMS [9]. These findings suggest that there are FMS patient subgroups with potentially distinct pathophysiological mechanisms at play, which may also be reflected by diversity in symptom manifestation [4, 10].

One of the most substantiated hypothesis on the pathophysiology of FMS is that of central sensitization. Central sensitization is defined as an amplification of neural signaling within the CNS that leads to pain hypersensitivity. However, it is unclear whether central sensitization happens primarily (“top-down”) or is a secondary consequence of peripheral nociceptive input (“bottom-up”) [1]. A primary central sensitization, which might lead to reduced descending pain inhibition, is supported by findings in the serotonergic-adrenergic [11] and opioid systems [12] in FMS patients. The excitatory neurotransmitter glutamate was investigated in context with central sensitization. Indicators are higher glutamate levels in cerebrospinal fluid of FMS patients compared to healthy controls or rheumatoid arthritis patients [11] and in MRI spectroscopy findings in FMS patients compared to healthy controls, especially of the insular cortex [13]. The clinical efficacy of memantine, an NMDA receptor antagonist, in alleviating pain [14], also points to a potentially important role of glutamate in the pathophysiology of FMS. Indicators of peripherally triggered sensitization in FMS are, for example, findings from microneurography, showing spontaneous activity and sensitization in C-fibers [15].

In the peripheral nervous system, damage to small nerve fibers has been shown in a clinically more severely affected subgroup of FMS patients [4, 16]. In an experimental animal study in rats, elevated glutamate levels in the insula led to a decrease in intraepidermal nerve fiber density (IENFD) [17].

The insular cortex is involved in different dimensions of pain [18, 19]. It is assumed to play an important role in the affective, cognitive, and sensory-discriminative aspect of chronic pain [20-22]. Previous neuroimaging studies discovered stronger insular connectivity to other pain processing brain areas through fMRI studies and higher glutamate levels in chronic pain disorders through magnetic resonance spectroscopy [23, 24]. As the anterior and posterior insular cortex have different functional connectomes to cortical and subcortical regions, both insular regions are assumed to perform different tasks in the processing of pain signals [25].

The anterior insular cortex is mainly involved in affective processing of pain stimuli and interospective awareness [26, 27]. In a magnetic resonance spectroscopy (MRS) study, low GABA levels were found in the anterior insular cortex of FMS patients compared to healthy controls. Electro acupuncture treatment of FMS patients was associated with an increased concentration of these GABA levels in the anterior insular cortex, which was associated with the subjective report of patients’ pain intensity [28].

The posterior insular cortex is considered a potential “gateway” to the somatosensory network. Focal epilepsies in this area may trigger pain sensations [29]. A MRS study showed higher glutamate levels in the right posterior insular cortex of FMS patients compared to healthy controls [13]. Lower GABA levels in the posterior insular cortex correlated with lower pressure-pain thresholds [30].

Most MRS studies investigated small groups of patients or did not compare findings with healthy and/or disease controls. Since CNS alterations reported in FMS were not specific, such comparisons are crucial. This is particularly true when assessing relative levels of neurotransmitters. Hence, in order to enable a direct comparison, this study recruited a large cohort of FMS patients, RA disease controls and healthy controls. We aimed to test the hypothesis that FMS patients have higher glutamate or lower GABA concentrations in the insular cortex, respectively. We further aimed to test the hypothesis that this mismatch is related to symptom severity. In an exploratory analysis, we further tested the hypothesis that higher glutamate levels are associated with a lower skin innervation in FMS patients.

## Materials and Methods

### Study design

In this case-control study, we recruited three groups of study participants: 1. A patient group with a diagnosis of FMS. 2. A patient group matched for age, sex and BMI with a diagnosis of RA as a disease control group. 3. A healthy control group matched for age, sex, and BMI. All groups received a single MRS examination. The patient groups (1, 2) also filled in clinical questionnaires.

### Participants

Patients were eligible if they were female, with an age range from 40 to 65 years, and a body-mass-index (BMI) between 20 and 35. Patients diagnosed with FMS met the diagnostic criteria according to the 2010 guidelines by the American College of Rheumatology [31]. MRI and clinical data of the FMS patients have already been published [4, 10]. RA patients (disease controls) were diagnosed according to the American College of Rheumatology/European League against Rheumatism (ACR/EULAR) classification criteria and had a disease duration of at least 5 years [32] with a respectively long chronic pain history. To assure ongoing disease activity, we only included RA patients with a disease activity score (DAS28) [33] ≥ 2.6 (exclusion of RA patients in remission) and clinically significant pain (minimum pain intensity on a numeric rating scale ≥ 4). No RA patient fulfilled the diagnostic criteria for FMS. The reason for including RA patients as disease controls was their persistent pain over several years, but with a well-defined inflammatory cause. The control group consisted of healthy women, also matched by age and BMI.

Patients and controls were excluded if they had any form of MRI incompatibility, other severe pain syndromes, other inflammatory diseases, any other neurological, psychiatric or severe cardiovascular diseases, history of drug abuse, head trauma requiring medical attention, or any structural anomaly of the brain. Written informed consent was obtained from all patients and controls. The ethics committee of the University of Würzburg University provided approval of the study (105/19).

### Clinical evaluation

In all patients, we evaluated pain intensity and pain-related disability through the Graded chronic pain scale (GCPS) [34] and used the German version of the center for epidemiological studies – Depression scale questionnaire (“Allgemeine Depressionsskala” (ADS)) [35] to record depressive symptoms. We further applied the “Pain Catastrophizing scale” [36]. We evaluated daily life impairment due to pain in all patients with the Fibromyalgia Impact Questionnaire (FIQ) [37]. All questionnaires are self-report measures. Disease activity of RA patients was assed using the DAS28, which includes the examination of swelling and pressure pain of 28 joints and the one hour erythrocyte sedimentation rate (ESR) [33].

### Evaluation of intraepidermal small fiber density

As described and published previously [4], FMS patients were evaluated for IENFD by skin punch biopsy at two sites (lower and upper leg). Based on the results, patients were divided into subgroups [10] with normal and reduced IENFD.

### MRS data acquisition

Magnetic resonance scanning was conducted on a 3T MR Scanner (Prisma, Siemens, Germany) with a 64-channel head coil at the Institute of Neuroradiology, University Hospital of Würzburg. We used a “MPRAGE” sequence to acquire T1-weighted anatomical images for spectroscopic voxel placement (repetition time TR = 2400ms, echo time (TE) = 3.17ms, flip angle (FA) = 8° inversion recovery (IR) = 1000ms, 176 sagittal slices, isotropic voxel size = 1 × 1 × 1 mm). MR-Spectroscopy is a non-invasive in vivo method to measure metabolites in the brain. GABA has proven to be difficult to detect due to its spectral peaks partially overlaping with peaks of other major metabolites especially creatinine (Cr), which is present in much greater concentrations. However, the use of editing sequences like MEshcher-Garwood Point-RESolved Single-voxel Spectroscopy (MEGA-PRESS) increased the reliability and is commonly used for GABA acquisition [38-42]. Therefore, a MEGA-PRESS sequence was used to detect GABA, glutamate and N-acetyl-aspartate (NAA) in the regions of interest (ROIs), with the following scanning parameters: TR = 2500 ms, TE = 68 ms, acquisition bandwidth = 1500 Hz, editing pulses frequency applied (off) at 1,9 ppm and (off) at 7.5 ppm, editing pulse bandwidth = 52 Hz. The 30 × 15 × 20 mm voxel was placed manually successively in the right and left IC. The voxel size was chosen as compromise between localization and signal quality to compensate the low signal to noise ratio (SNR) for GABA (0.7 to 1.4 mM/cm3) [39, 43]. To compromise between SNR (thus reliable results) and anatomical specificity, the entire insular cortex was used as voxel content rather than the subdivision into anterior and posterior. Before the measurement, the magnetic field was automatically shimmed in 3 rounds on the respective voxel. Only measurements with magnetic field inhomogeneity <30 Hz average linewidth were accepted for the defined ROI. An unsuppressed water measurement was used for frequency and phase correction and as reference to tissue water.

### Spectroscopy analysis

Preprocessing and fitting of the MR spectra was performed using the Totally Automatic Robust QUantitation in Nuclear MR (TARQUIN) software. The metabolite concentrations of GABA, Glutamate and NAA were analyzed. Glutamate concentrations were obtained from the off spectra. GABA and NAA concentrations were obtained from the difference spectrum (on – off).

### Statistical Analysis

Statistics were performed with JASP Team (2022). JASP (Version 0.16.2) [Computer software]. The data were tested for normal distribution with a Shapiro-Wilk test. We used the Levene test with a significance threshold of 0.05 % to check the data for equivalence of variance. To compare the groups to each other regarding the concentration of the metabolites and the clinical parameters, we calculated ANOVAs. Due to the different group sizes, a Welch correction was performed and the following post-hoc comparisons were corrected for multiple comparisons using the Games-Howell method. Depending on the condition of normal distribution, correlation analyses were calculated using Pearson’s or Spearman’s correlation. Due to higher standard deviations, we calculated subgroup comparisons using the Mann-Whitney-U Test. The confidence interval was 95 %, the significance level at 0.05 %.

## Results

### Cohort characteristics

The final sample size consisted of 102 participants (all female), who were divided into three groups: FMS group (n = 44; subgroup with reduced IENFD (PNS) n = 21; subgroup with normal IENFD (noPNS) n = 23), RA group (n = 19), and healthy controls (n = 40). FMS patients showed higher scores than RA patients in the self-report questionnaires on pain intensity, impairment in daily life, depression and pain catastrophizing. The group differences in clinical symptoms and structural and functional imaging between the FMS group and healthy controls have already been published [10]. All baseline characteristics can be found in Table 1.

**Table 1:**
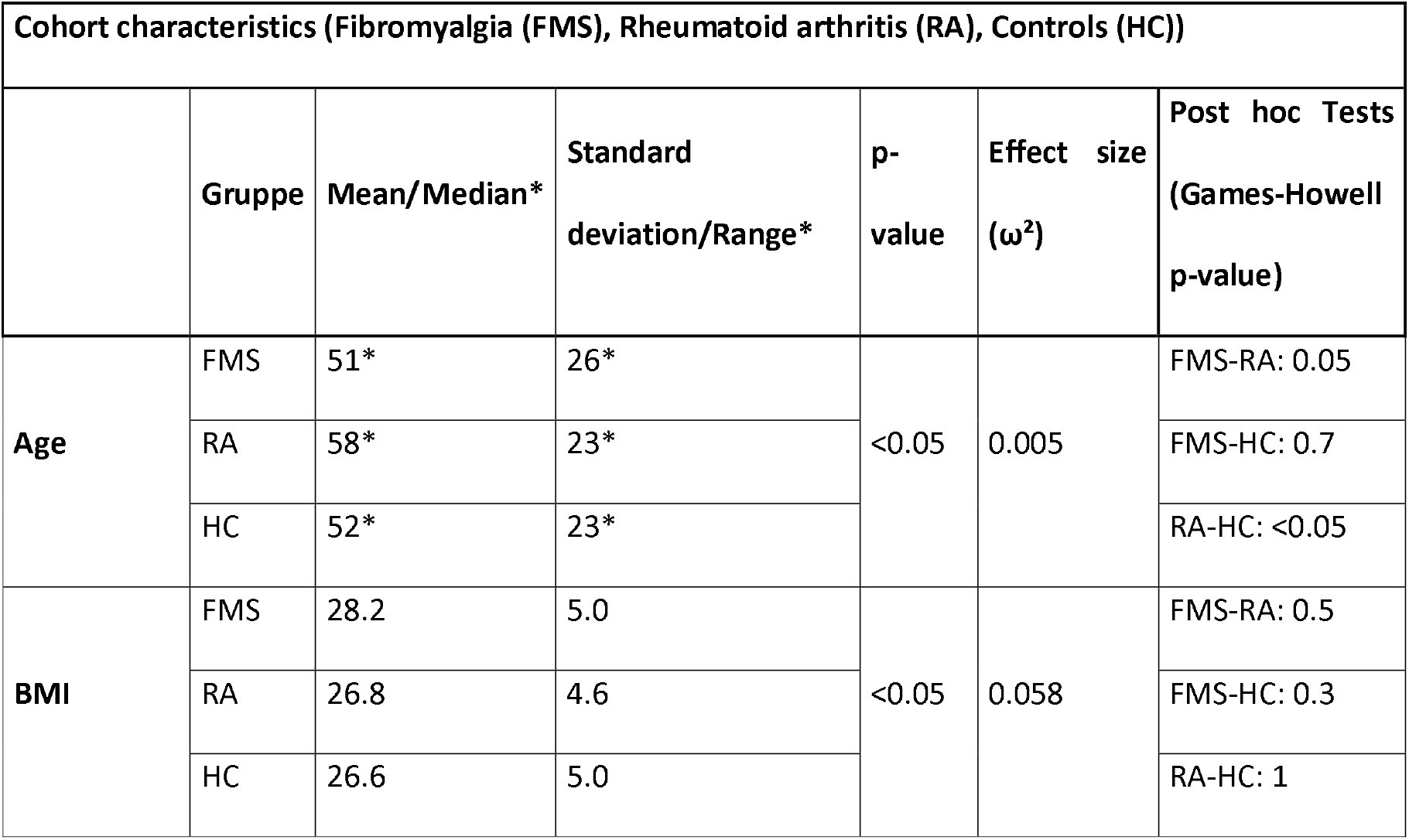
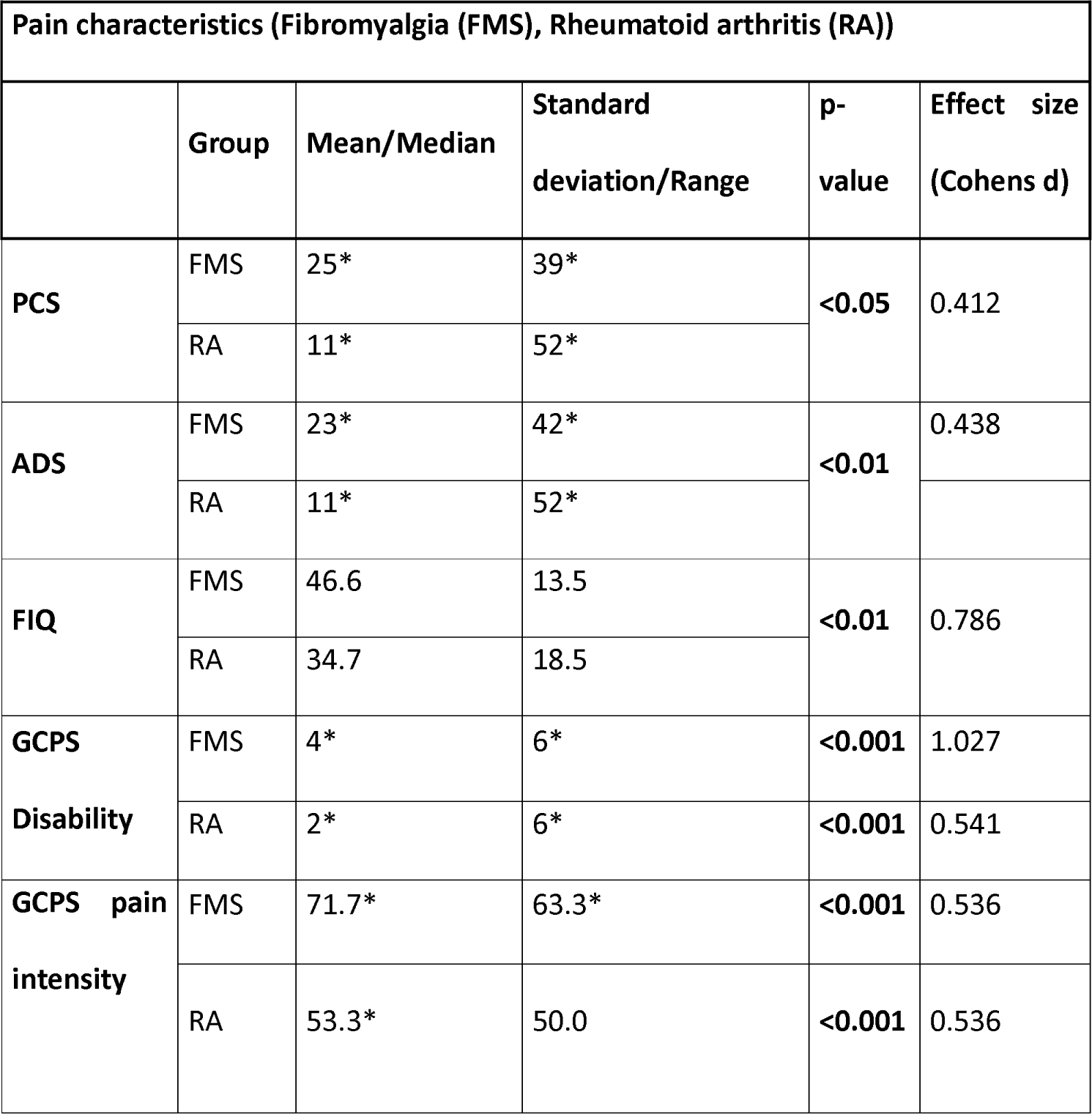
Group characteristics. BMI: Body Mass Index, FMS: Fibromyalgia-syndrome, RA: Rheumatoid arthritis, PCS: Pain catastrophizing scale (sum score), ADS: Allgemeine Depressionsskala, FIQ: Fibromyalgia Impact Questionnaire, GCPS: Graded chronic pain scale. *This variable was not normally distributed. The data are therefore characterized as median/range.

### Group differences in glutamate, GABA, and NAA

Glutamate and GABA concentrations did not differ in the insular cortex between the FMS and the control groups. Only the concentration of NAA in the insular cortex of RA patients was lower compared to the FMS group (left hemisphere p-value: 0.08, right hemisphere p-value: 0.003) and the control group (left hemisphere p-value: 0.02, right hemisphere p-value: 0.02) (Figure 1, Table 2).

**Figure 1:**
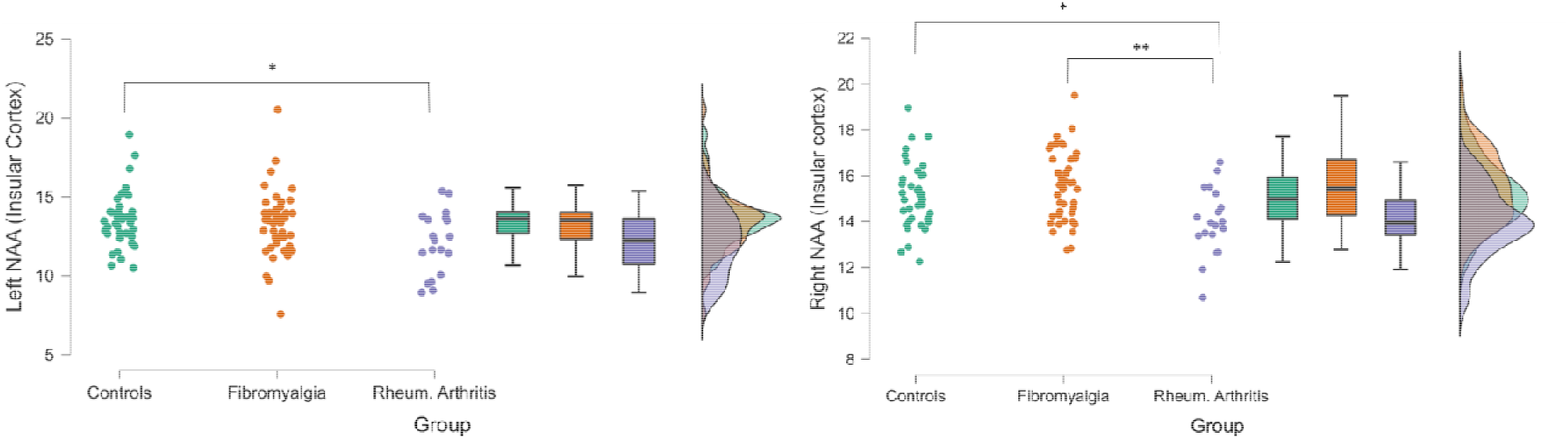
Lower NAA concentration of the insular cortex of RA patients compared with controls (left and right hemisphere) and with fibromyalgia patients (right hemisphere).

**Table 2:**
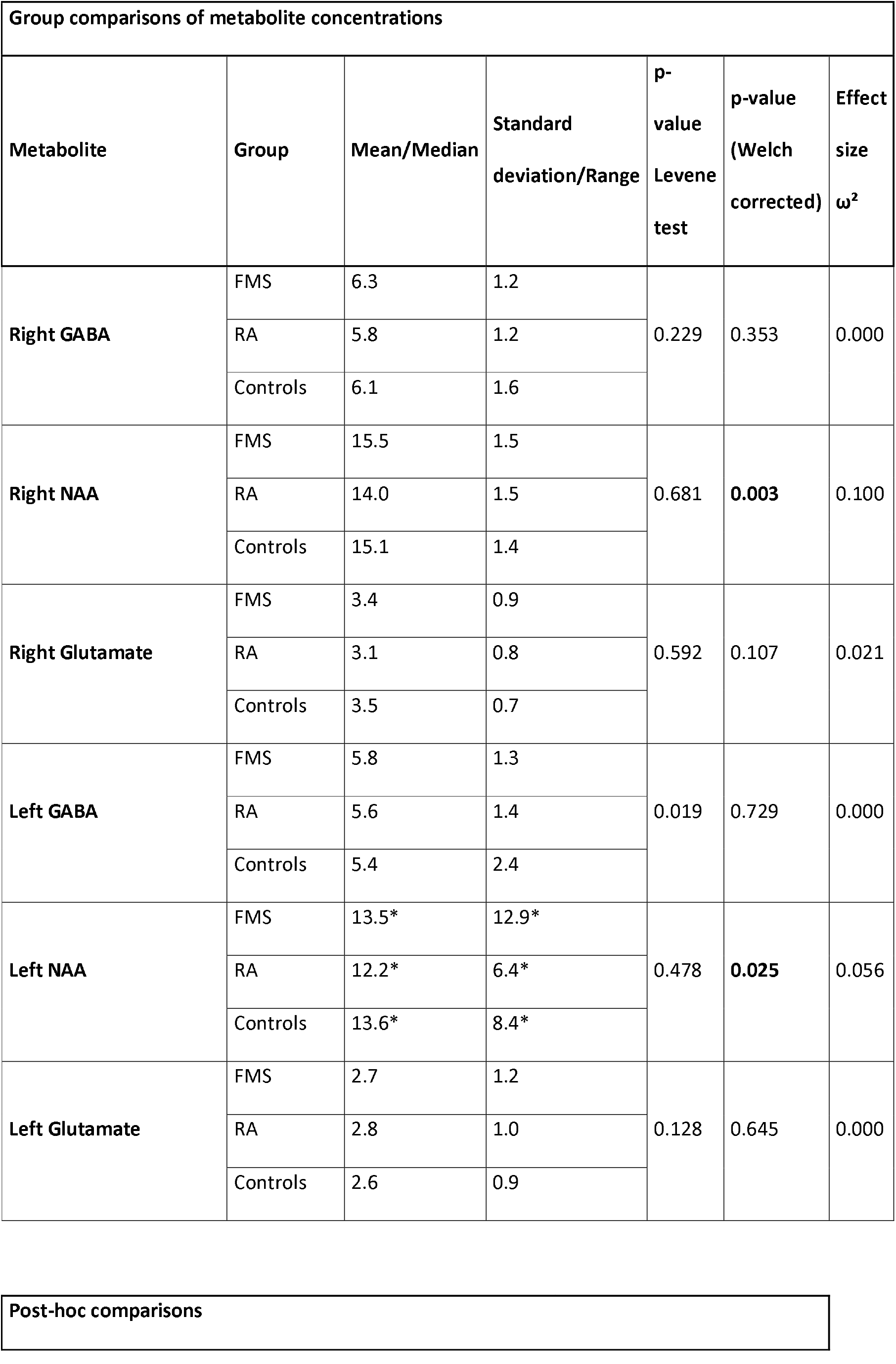
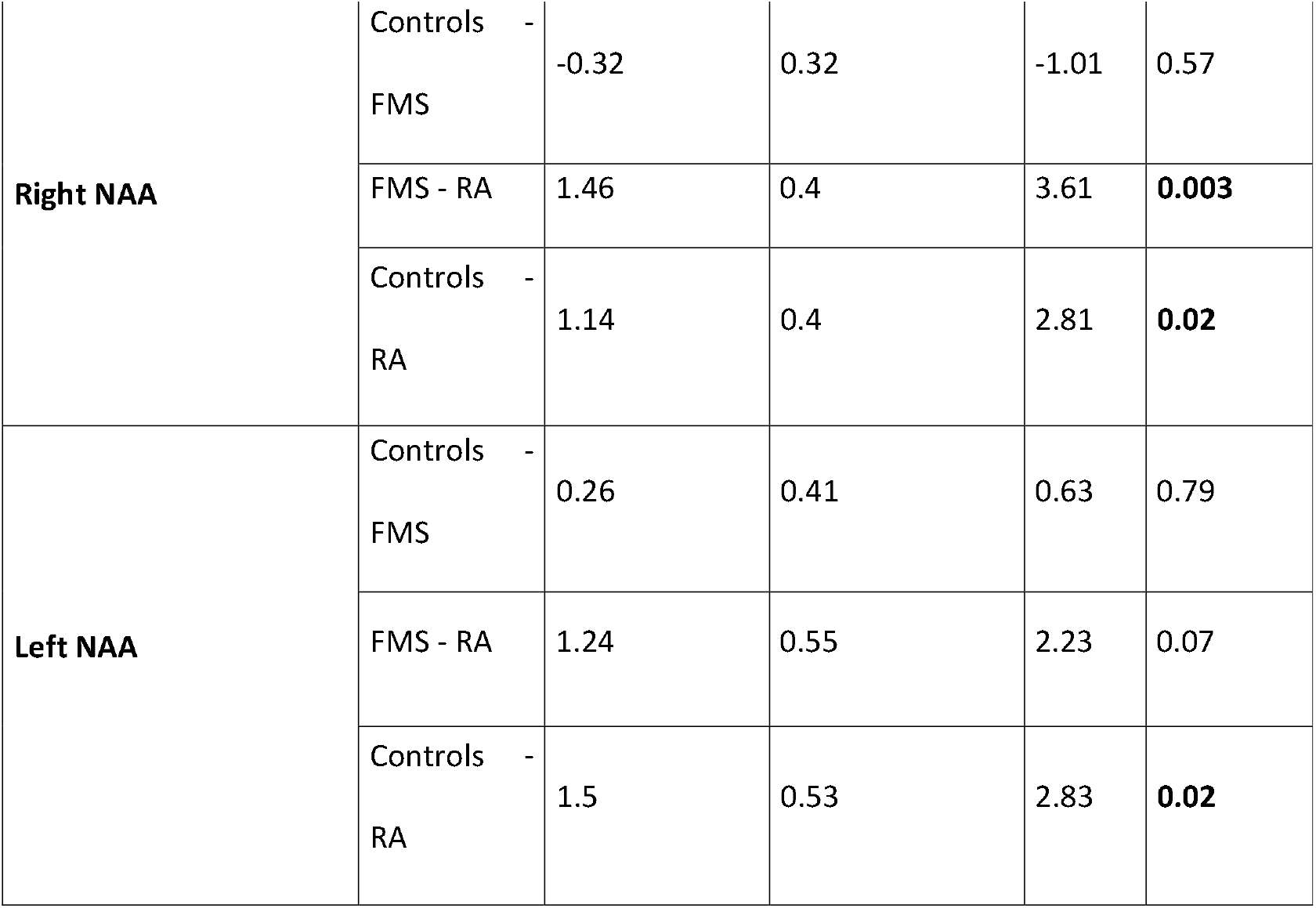
Group comparisons of metabolite concentrations in the left and right insular cortex. NAA: N-acetylasparate, GABA (gamma-amino-butyric acid), FMS: Fibromyalgia syndrome, RA: Rheumatoid arthritis. *This variable was not normally distributed. The data are therefore characterized as median/range.

| Group comparisons of metabolite concentrations |  |  |  |  |  |  |
| --- | --- | --- | --- | --- | --- | --- |
| Metabolite | Group | Mean/Median | Standard deviation/Range | p-value<br>Levene test | p-value<br>(Welch corrected) | Effect size<br>$\omega^2$ |
| Right GABA | FMS | 6.3 | 1.2 | 0.229 | 0.353 | 0.000 |
|  | RA | 5.8 | 1.2 |  |  |  |
|  | Controls | 6.1 | 1.6 |  |  |  |
| Right NAA | FMS | 15.5 | 1.5 | 0.681 | 0.003 | 0.100 |
|  | RA | 14.0 | 1.5 |  |  |  |
|  | Controls | 15.1 | 1.4 |  |  |  |
| Right Glutamate | FMS | 3.4 | 0.9 | 0.592 | 0.107 | 0.021 |
|  | RA | 3.1 | 0.8 |  |  |  |
|  | Controls | 3.5 | 0.7 |  |  |  |
| Left GABA | FMS | 5.8 | 1.3 | 0.019 | 0.729 | 0.000 |
|  | RA | 5.6 | 1.4 |  |  |  |
|  | Controls | 5.4 | 2.4 |  |  |  |
| Left NAA | FMS | 13.5* | 12.9* | 0.478 | 0.025 | 0.056 |
|  | RA | 12.2* | 6.4* |  |  |  |
|  | Controls | 13.6* | 8.4* |  |  |  |
| Left Glutamate | FMS | 2.7 | 1.2 | 0.128 | 0.645 | 0.000 |
|  | RA | 2.8 | 1.0 |  |  |  |
|  | Controls | 2.6 | 0.9 |  |  |  |

|  |
| --- |
| Post-hoc comparisons |

Table 2: Group comparisons of metabolite concentrations in the left and right insular cortex.
| Metabolite | Group comparison | Mean Difference | Standard error | t-value | p-value (Games Howell corrected) |
| --- | --- | --- | --- | --- | --- |
| Right NAA | Controls - FMS | -0.32 | 0.32 | -1.01 | 0.57 |
|  | FMS - RA | 1.46 | 0.4 | 3.61 | **0.003** |
|  | Controls - RA | 1.14 | 0.4 | 2.81 | **0.02** |
| Left NAA | Controls - FMS | 0.26 | 0.41 | 0.63 | 0.79 |
|  | FMS - RA | 1.24 | 0.55 | 2.23 | 0.07 |
|  | Controls - RA | 1.5 | 0.53 | 2.83 | **0.02** |

### Subgroup comparisons

Subgroups with normal IENFD (noPNS; n=23) and reduced IENFD (PNS; n=21) did not differ in metabolite concentrations (Table 3).

**Table 3:** Comparison of metabolite concentrations in the subgroups PNS (reduced IENFD) and noPNS (normal IENFD).

| Subgroup metabolite concentrations |  |  |  |  |  |
| --- | --- | --- | --- | --- | --- |
| Metabolite | Subgroup | Median | Range | p-value | Cohen's d |
| Right GABA | noPNS | 8.6 | 3.2 | 0.38 | 0.16 |
|  | PNS | 6.0 | 4.5 |  |  |
| Right NAA | noPNS | 15.6 | 5.3 | 0.9 | 0.01 |
|  | PNS | 15.1 | 5.7 |  |  |
| Right Glutamate | noPNS | 3.6 | 4.2 | 0.78 | 0.05 |
|  | PNS | 3.6 | 3.2 |  |  |
| Left GABA | noPNS | 5.7 | 4.0 | 0.59 | 0.09 |
|  | PNS | 5.7 | 4.9 |  |  |
| Left NAA | noPNS | 13.6 | 12.9 | 0.85 | -0.03 |
|  | PNS | 13.3 | 6.0 |  |  |
| Left Glutamate | noPNS | 2.4 | 4.9 | 0.59 | -0.09 |
|  | PNS | 3.0 | 4.7 |  |  |

### Associations of clinical symptom severity and cerebral levels of glutamate and GABA

We found no evidence that an increased insular glutamate concentration or decreased GABA concentration in FMS patients are related to more severe clinical symptoms. We found no correlations in the FMS group nor in the patient group (RA + FMS patients combined). More detailed statistics on “metabolite ∼ clinical parameter” associations in FMS patients can be found in Table 4.

**Table 4:** Associations with metabolite concentrations and the severity of clinical parameters of FMS Patients. PCS: Pain catastrophizing scale (sum score), ADS: Allgemeine Depressionsskala, FIQ: Fibromyalgia Impact Questionnaire, GCPS: Graded chronic pain scale.

| Clinical parameter |  | Right<br>Glutamate | Right GABA | Left<br>Glutamate | Left GABA |
| --- | --- | --- | --- | --- | --- |
| **PCS** | Spearman's<br>rho | -0.089 | 0.054 | -0.114 | -0.003 |
|  | p-value | 0.572 | 0.732 | 0.465 | 0.984 |
| **ADS** | Spearman's<br>rho | 0.148 | -0.125 | 0.164 | -0.110 |
|  | p-value | 0.338 | 0.418 | 0.288 | 0.476 |
| **FIQ** | Spearman's<br>rho | 0.040 | -0.178 | 0.224 | 0.002 |
|  | p-value | 0.795 | 0.247 | 0.144 | 0.992 |
| **GCPS Pain intensity** | Spearman's<br>rho | -0.262 | 0.077 | -0.078 | 0.040 |
|  | p-value | 0.086 | 0.621 | 0.614 | 0.795 |
| **GCPS Disability due**<br>**to pain** | Spearman's<br>rho | -0.049 | -0.065 | -0.074 | 0.053 |
|  | p-value | 0.754 | 0.674 | 0.635 | 0.731 |

## Discussion

In this study including 102 participants, we found no evidence for higher glutamate or lower GABA levels in the insular cortex of FMS patients compared with healthy and disease controls. We also found no evidence that higher levels of glutamate correlated with lower IENFD. The levels of glutamate and GABA in the insular cortex did not correlate with the severity of classic symptoms in FMS patients in our analysis.

The utility of non-invasive MRS as a method to better explore the pathophysiology of chronic pain disorders has increased [44, 45]. In addition to measuring glutamate, there are also some studies that have found decreased levels of NAA in various brain regions of FMS patients, such as the dorsolateral prefrontal cortex [46] and the hippocampus [47]. Higher concentrations of Glx (glutamine + glutamate) were also found in other regions such as the posterior gyrus of 10 FMS patients compared to 10 healthy controls [48]. Regarding the target region of the present study, the insular cortex, the body of evidence for a static change in glutamate/GABA changes is relatively small. An MRS study of 19 FMS patients and 14 controls found an increase in glutamate in the right posterior insular cortex in FMS, which correlated with decreased pain pressure thresholds [13]. A few years later, in a study of 16 FMS patients and 17 controls, the same research group published decreased GABA concentrations in the anterior right insular cortex but not in the posterior. However, in the posterior insular cortex of FMS patients, GABA levels still correlated with the pressure pain threshold [30]. A PET study suggested that the increased GABAa receptor density in different brain regions in FMS patients might be a compensation for the decreased GABA concentrations [49]. In recent years, the focus has shifted from static differences in glutamate/GABA concentrations to dynamic concentrations after interventions. A combined MRS and fMRI study with 67 FMS patients but no controls showed that electrical acupuncture could elicit pain relief in FMS patients. This pain relief correlated with increased connectivity of the anterior insular cortex and a part of the somatosensory cortex. The connectivity strength in turn correlated with an increase in GABA in the anterior insular cortex [28]. A similar study with 17 FMS patients, also without controls, showed that the administration of pregabalin alleviated FMS pain. This pain relief correlated with a reduced connectivity of the anterior insular cortex with the default mode network. This reduced connectivity, in turn, correlated with a decreased Glx concentration in the posterior insular cortex [50]. Thus, there is some evidence that suggests a mismatch of glutamate/GABA in the insular cortex of FMS patients. The glutamate/GABA mismatch seems to correlate with the severity of clinical symptoms, however, the sample size was small.

Our finding that RA patients show decreased levels of NAA, a marker of neuronal integrity [51], has already been supported in the literature regarding RA and other rheumatic diseases [52]. The most consistent conjecture on the background of decreased NAA concentrations in the magnetic resonance spectroscopy literature on pain is that of neuronal damage. Decreased NAA levels also appear to be related to increased inflammatory markers. [53]. Therefore, the normal NAA concentrations in our study in FMS patients may indicate intact neuronal integrity of the insular cortex. Only in one brain region, the hippocampus, a meta-analysis found decreased NAA levels FMS patients. [47].

There are several potential reasons why our study did not report group differences, which is not in line with previous findings on a glutamate/GABA mismatch of the insular cortex in FMS patients: Most of the previous studies had subdivided the insular cortex into the anterior and posterior insular cortex as the regions of interest (ROI) [13, 30, 50]. Splitting the insular cortex in two subregions is reasonable, as there is evidence from fMRI studies that the anterior and posterior insular cortex perform different functions in pain processing [22]. However, in planning the study design, we had opted to measure the entire insular cortex for several reasons. One reason was that previous literature has been incongruent as to whether the glutamate/GABA mismatch is present in the anterior or posterior insular cortex [13, 30]. Another reason for measuring the entire insular cortex was that MEGA-PRESS sequences have a particularly unfavorable signal to noise ratios [43].

In a recent MRS study increased GABA concentrations were found in FMS patients in the anterior insula after electronic acupuncture [28]. Administration of pregabalin decreased glutamate concentrations in the posterior insula of 17 FMS patients [50]. Decreased connectivity with the default mode network then correlated with decreased expression of FMS symptoms. However, because our study used a static setting to measure metabolite concentrations and the aforementioned dynamic studies did not include a comparison with healthy controls in their study design, no comparisons can be made here.

Finally, MEGA-PRESS sequence are susceptible to confounding and therefore do not only depend on physical prerequisites, such as the correct shimming for homogenization of the magnetic field lines and the voxel size, but also on the analysis method [43]. It has been shown that the standard softwares in MRS analysis differ in their technique of fitting the spectra, resulting in different quantification results of the MEGA-PRESS metabolite concentrations [54].

Our study has several limitations. Because of the measurement of the entire insular cortex, a subdivision into anterior and posterior insular cortex and a direct comparability with previous studies is only possible to a limited extent. Furthermore, due to the overlapping spectra of glutamine and glutamate, it is currently technically impossible to completely differentiate these two metabolites in MRS. Different analysis software may do the fitting of these two metabolite spectra to different extents. Another issue that may generate possible variations in metabolite concentrations is the subjective ROI placement on the insular cortex directly at the scanner. Because the anatomy of the insular cortex is individual, the ROI was placed directly at the scanner to allow the highest possible proportion of gray matter in the ROI. In addition, many cell types are also present in the gray matter besides neurons. Since glutamate is also a product of metabolism in, for example, glial cells, and not only a neurotransmitter of the synapses, this study describes metabolites rather than neurotransmitters concentrations. A final limitation is that measurements were performed at only one time point. A longitudinal study design could account for possible individual variations in metabolite concentrations in the future.

## Conclusion

Our study found no differences of glutamate/GABA concentrations between FMS and RA patients, nor healthy controls. Our data therefore does not substantiate the glutamate/GABA hypothesis of the insular cortex in the pathophysiology of FMS, which has been previously described in the literature. For the future, it would be beneficial to find uniform measurement methods for the quantification of the metabolites glutamate/GABA in order to compare them more directly. In addition, it would be important to conduct further longitudinal studies that could detect a potential causal effect on the glutamate/GABA metabolite concentrations with the help of an intervention.

## Data Availability

All data produced in the present study are available upon reasonable request to the authors.

## Acknowledgements

We would like to acknowledge Edward J. Auerbach, Ph.D. and Małgorzata Marjańska, Ph.D. (Center for Magnetic Resonance Research and Department of Radiology, University of Minnesota, USA) for the development of the pulse sequences for the Siemens platform which were provided by the University of Minnesota under a C2P agreement.

